# Variation in English Covid booster uptake

**DOI:** 10.1101/2022.02.01.22270236

**Authors:** Greg Dropkin

## Abstract

**Introduction:** Variable and low takeup of the Covid booster is a recognised problem, associated with age, gender, ethnicity, and deprivation. Are there other relevant predictors?

**Methods:** Data was downloaded from the UK Government Coronavirus Dashboard for Middle Super Output Areas in England, along with demographic, employment, and health data from public sources. Mixed models with a random factor for Upper Tier Local Authority were analysed as quasibinomial Generalized Additive Models, and the estimated random factors were then fitted with Bayesian linear mixed models using flu vaccination uptake, change in public health budgets, vaccination centres, and Region.

**Results:** Models for the MSOA Covid 1^st^ and 2^nd^ vaccinations and the 3^rd^ injection (including the booster), fit the data well. Index of Multiple Deprivation, proportion Aged 15-24 and 25-44, and ethnicity groupings Other White, Indian-Pakistani-Bangladeshi, and African-Caribbean-Other, are highly significant predictors of lower uptake. The estimated random factors vary widely amongst local authorities, and can be predicted by flu vaccine uptake, rise in public health budgets, and regional effects which are positive for London and South East, and negative for North West and North East. Vaccination centres did not reach 90% significance.

**Discussion:** Covid vaccination rates at each stage are very well modelled if local authority random effects are included along with non-linear terms for demographic, employment and health data. Deprivation, younger age, and Other White, South Asian, and Afro-Caribbean ethnicities are associated with lower uptake. Modelling the local effects indicates that increasing public health budgets would improve vaccination uptake.

## Introduction

The booster programme has been central to the UK government’s strategy for containing Covid-19 in England during the autumn and winter of 2021-22.^1^ However, takeup of the booster remains well below the levels achieved for the first and second vaccine doses.^2^ Vaccination uptake is highly dependent on age, gender, ethnicity, and deprivation, as is widely recognised ^3^ ^4^ ^5^ ^6^ ^7^ ^8^ ^9^

Demography varies across England, so vaccination rates will also vary. For example, as the Omicron variant swept the country on 4 January, the cumulative Third Injection (Booster or 3^rd^ primary) uptake in Newham was 27.2%, in contrast with 41.1% in Liverpool and 67.4% in Gloucestershire. These are different populations, but does demography explain the disparity in full? If there are additional sources of variation, are they characteristics of people or of where they live?

I explore these questions by modelling cumulative Covid vaccination in 6789 Middle Super Output Areas of England, each comprising around 8,000 people on average. The models use demographic, employment and health data at MSOA level and random effects for the 149 Upper Tier Local Authorities. The resulting estimated random effects are then modelled with selected predictors available at UTLA level: Region, flu vaccination rate, the number of vaccination centres, and annual change in local authority public health budgets. Regional disparity, public experience and attitudes to vaccination in general, access to Covid-19 vaccines, and public health department capacity, are all plausible influences on how local authorities may differ even while controlling for other factors.

## Data sources

The cumulative number of persons vaccinated, separately for each of two doses and the Third Injection, is available daily at MSOA level from the UK Coronavirus Dashboard ^10^ along with the Vaccine Register Population. The Third Injection comprises people given the Booster plus people over 12 with severely weakened immune system given a 3^rd^ (primary) dose. The Dashboard also provides MSOA incidence (“7 day rolling rate”) of Covid cases, weekly through to 5 days before download. Dashboard data was downloaded on 4 January 2022.

The Index of Multiple Deprivation (IMD), and separate indices for the Health and Education deprivation domains, were taken from the English indices of deprivation 2019 ^11^ using File 7: all ranks, deciles and scores for the indices of deprivation, and population denominators. Lower Super Output Area population and area (mid-2019 estimates) ^12^ were used to obtain density at MSOA level. Population by MSOA was taken from the mid-2019 estimates ^13^ which include data by gender and each year of age. Ethnicity at MSOA level from the 2011 Census was downloaded from NOMIS for the dataset DC2101EW - Ethnic group by sex by age ^14^. Employment by industrial sector is available at MSOA level from the 2011 Census and was downloaded from NOMIS dataset DC6110EW ^15^. Communal establishments data is available at MSOA level from the 2011 Census and was downloaded from NOMIS dataset QS421EW ^16^. Multi-occupation housing is available at MSOA level from the 2011 Census and was downloaded from NOMIS dataset DC1109EW ^17^.

Mean distance to GP, Emergency Department, and Pharmacy is available at LSOA level from the AHAH dataset ^18^. GP registration mapped to LSOA is available from NHS Digital ^19^. A lookup from MSOA to Local Authority to Upper Tier Local Authority to Region was available from Public Health England as part of weekly surveillance. The 28 October 2020 report is online, with lookup in columns 1-8 (select all, show columns) ^20^.

Seasonal Flu Vaccine Uptake data for 2020/21 was released by Public Health England ^21^. Data was extracted for GP-registered patients aged under 65, by Local Authority. The uptake for “Leicester and Rutland” was then assigned to each area separately. Covid-19 Vaccination Centres as of 17 November 2021 were listed by NHS England ^22^. Their postcodes were mapped to LTLA codes with the ONS National Statistics Postcode Lookup ^23^, and then to UTLA codes via the lookup above. Public Health ring-fenced grant allocations for 2020/21 and 2021/22 are available from the Department of Health ^24^.

## Statistical method and models

The first stage of modelling seeks to explain the observed vaccination rates in terms of demographic, employment and health data at MSOA level along with random effects for the 149 UTLA.

Separately for each of three vaccinations, cumulative numbers of persons vaccinated were combined with the Vaccine Register Population to form a matrix **vmat** with the numbers vaccinated and unvaccinated in each of 6789 MSOA, as of 4 January 2022. For each vaccination, the corresponding matrix (**vmat1** etc) is the response variable. Four models were considered, differing in the choice of explanatory covariates, and fitted separately for each vaccination.

The first stage models were structured as Generalized Additive Models, using the R package “mgcv” ^25 26^. The factor **utla** with 149 levels denotes the UTLA for each MSOA, and is treated as a random effect. All other covariates are treated as fixed effects.

Covariates for IMD, cumulative incidence, and population density were scaled. Most other covariates were transformed to improve performance. For a covariate **v**, the monotonic transform **LV** is defined as log(1+**v**/mean(**v**)). For example, the population proportion for a particular ethnicity or industrial sector may be very low, but its transformation will be more easily smoothed.

The random effect smoother has the form s(utla, bs=“re”) whilst other smoothers have the form s(v, bs=“cr”, k=7) using cubic regression splines with basis dimension 7. Simplifying the notation from IMD onwards, the first model is

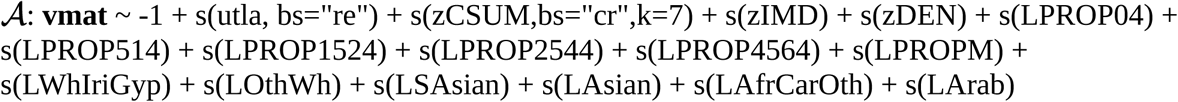

This model uses the random factor **utla**, cumulative incidence, IMD, population density, five age bands (as transformed proportions), male (transformed proportion), and six ethnicities (transformed proportion). The latter, using the designations from the 2011 Census, are White Irish and White Gypsy (combined group); Other White; Indian, Pakistani, and Bangladeshi (combined group); Chinese and Other Asian (combined group); African, Caribbean, Other Black, and Other Ethnicity (combined group); Arab. The initial -1 removes the intercept so that all levels of **utla** are treated equally.

Model ***ℬ***enlarges ***𝒜*** with additional terms for the transformed population proportions of 15 industrial sectors (Agriculture Energy Water, Manufacturing, …, Other)

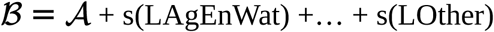

Model ***𝒞*** enlarges ***𝒜*** with additional demographic and local health covariates for Communal establishments, multi-occupation housing, population weighted mean distance to GP, to A&E, to Pharmacy, proportion registered with GP, Health deprivation, and Education deprivation

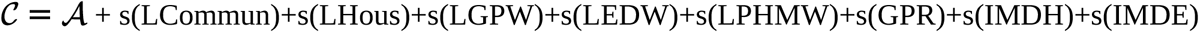

Model ***𝒟*** enlarges ***𝒜*** with most of the additional terms from ***ℬ*** and ***𝒞***, but omits six terms whose contributions showed negligible or low significance: Density, Manufacturing, Accommodation and Food, Information and Communication, Real Estate, mean distance to GP.

Models were fitted separately for each vaccination, using the quasibinomial family with smoothing parameter optimisation by marginal likelihood (method = “ML”). ^27^ The output includes the fitted model coefficients *b* and their Bayesian posterior covariance matrix *V*_*c*_ which includes correction for smoothing parameter uncertainty (an option with the “ML” method). Model fit was evaluated with gam.check, and outliers detected by cooks.distance > 0.02, a somewhat arbitrary criterion. The actual and predicted proportions vaccinated in each MSOA were plotted, along with the smoother curves for particular covariates. Models were compared by ML value (lower value indicates better fit).

For each fitted model, the first 149 coefficients c_i_ are the estimated contributions of the levels of **utla**. Each c_i_ has a standard error se_i_, obtained as the square root of the i^th^ diagonal term of *V*_*c*_. “Caterpillar plots” were drawn using the c_i_ and se_i_ to show the variation in estimated random effects.

These coefficients also enable comparison of the fitted value for a particular MSOA, with the hypothetical fitted value if the same population were located in a different UTLA, replacing c_i_ with c_j_. If g denotes the (quasi) binomial link g(p) = log(p/(1-p)) and h is the inverse link h(t) = 1/(1+exp(-t)), the fitted value would be altered from fit_1_ to h(g(fit_1_) - c_i_ + c_j_).

In a second stage of modelling, the c_i_ were taken as observed, to be predicted using covariates available for Upper Tier Local Authorities. Region was treated as a random effect, with fixed effects for the uptake of Flu vaccination, the change in Public Health ring-fenced grant allocations from 2020-21 to 2021-22, and the number of Covid Vaccination Centres within the local authority.

This stage used the Bayesian programme rstanarm ^28^ to obtain parameter estimates and credible intervals, and a plot of predicted and observed c_i_. Using the associated package “loo” ^29^, a pointwise value of pareto_k > 0.7 was taken to indicate an outlier.^30^ A Bayesian version of R^2^ is used to describe overall model fit.^31^

In fact the c_i_ are not observed, but are an output of first stage modelling with associated standard errors. To estimate the impact of this uncertainty, simulated **c**\* were drawn as multivariate normal with mean **c** (the vector with components c_i_) and variance *V*_*c*_. Second stage modelling was repeated using the simulated **c**\* to give fresh sampling output (4000 rows), and this process was repeated 100 times to produce a combined sampling matrix with 404,000 rows (including the original output from second stage modelling of the c_i_). The mean, 5% and 95% quantiles of its columns were taken as corrected estimates and credible intervals of the second stage parameters, taking into account the uncertainty in the c_i_.

## Results

ML values and proportion of Deviance explained for the 4 models and 3 doses are shown in Table 1

**Table 1:** ML value and proportion of Deviance explained.

| Model | Dose 1 | Dose 2 | 3<sup>rd</sup> Injection |
| --- | --- | --- | --- |
| *A* | -15385.47 (0.954) | -15974.68 (0.959) | -15669.84 (0.965) |
| *B* | -15743.80 (0.959) | -16259.34 (0.963) | -16066.13 (0.970) |
| *C* | -16032.83 (0.962) | -16560.01 (0.966) | -16031.49 (0.969) |
| *D* | -16243.52 (0.965) | -16746.87 (0.968) | -16278.81 (0.972) |

The fitted models pass gam.check. For each vaccination, 𝒟 has the lowest ML value and the highest proportion of Deviance explained.

For the 3^rd^ Injection with model 𝒟, all predictors have highly significant effects (p<0.001) except for distance to pharmacy (p<0.01). 𝒟 explains 97.2% of the deviance, whilst a model using only the random effects term explains 51.5%, and a model using only the fixed effects explains 95.7% (percentages do not sum to 97.2% as in the absence of some smoothers, others compensate). Both of the reduced models have considerably worse fit than 𝒟 (higher ML values), and are far inferior by anova tests.

Charts were produced for the 3^rd^ Injection with model 𝒟.

Chart 1 shows the observed and fitted values of uptake, the outliers with cooks.distance > 0.02 and the 37 MSOA within Newham.

**Chart 1:**
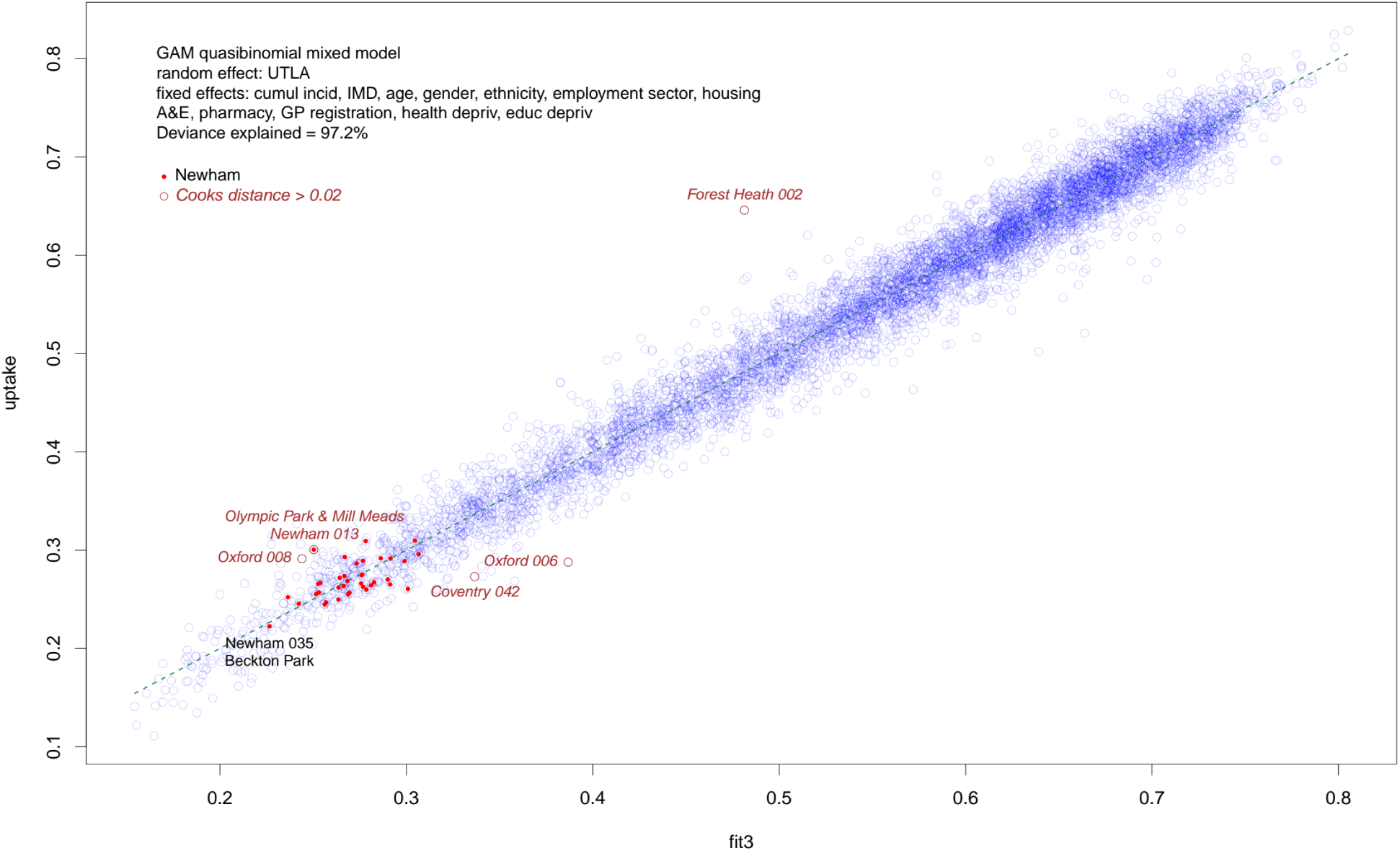
fitted value vs uptake Third Injection data as of 4 Jan 2022, model D.

Cooks distance is a metric combining the influence of individual points on the fitted model, and the magnitude of their deviance residual. There are only five outliers as judged by cooks.distance > 0.02, and of these only Forest Heath 002 (Lakenheath) has a high residual, the others being highly influential points. Newham 013 (Olympic Park & Mill Meads) is an outlier, with uptake higher than expected. Newham 035 (Beckton Park), with the lowest uptake (0.223) in this local authority, nearly matches its predicted value (0.227). Overall, the model fits the data very well.

Chart 2 shows selected smoothers. These plot the modelled impact of individual covariates on the overall fitted value (displayed on the scale of the linear predictor, before it is translated to fitted output by the inverse link function). The x-axis is limited by the 0.01 and 0.99 quantiles of the covariate. The first 6 smoothers shown have much larger impact on fitted values, and each of them has a negative effect: higher values of scaled IMD give lower fitted values, and likewise for the age bands and ethnicities shown. In the subsequent panels the y-axis is limited by the range of the smoother and is labelled “narrow scale”. Thus an increasing male proportion also has a negative impact, but its effect is smaller than that of IMD, age, or ethnicity.

**Chart 2:**
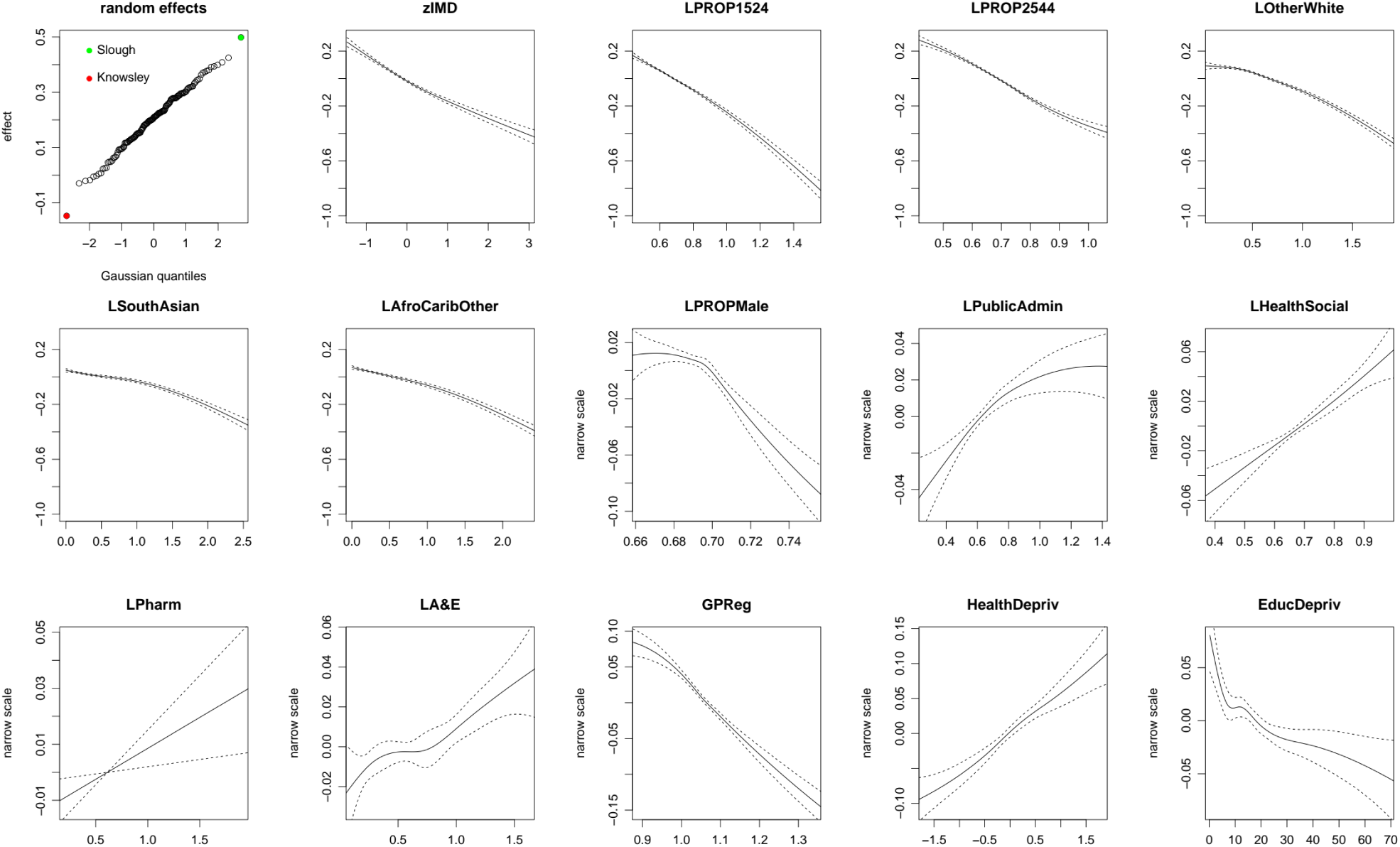
Selected smoothers for model *𝒟*.

Chart 3 shows the “Caterpillar plot” for the estimated random effects.

**Chart 3:**
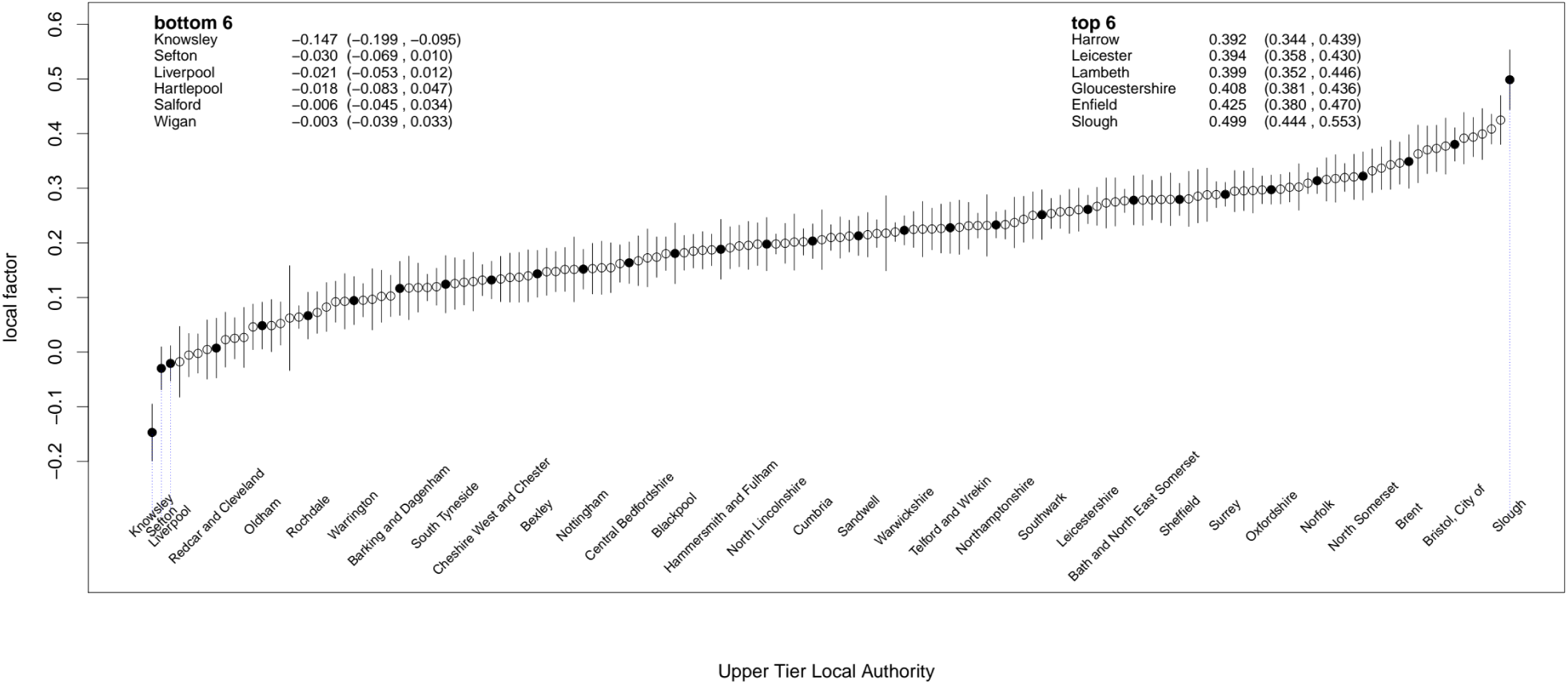
3rd injection local factors data as of 4 Jan 2022, model D.

Chart 4 shows the parameter estimates and 90% credible intervals from second stage modelling of the scaled estimated random effects from model ***𝒟*** for the 3^rd^ injection.

**Chart 4:**
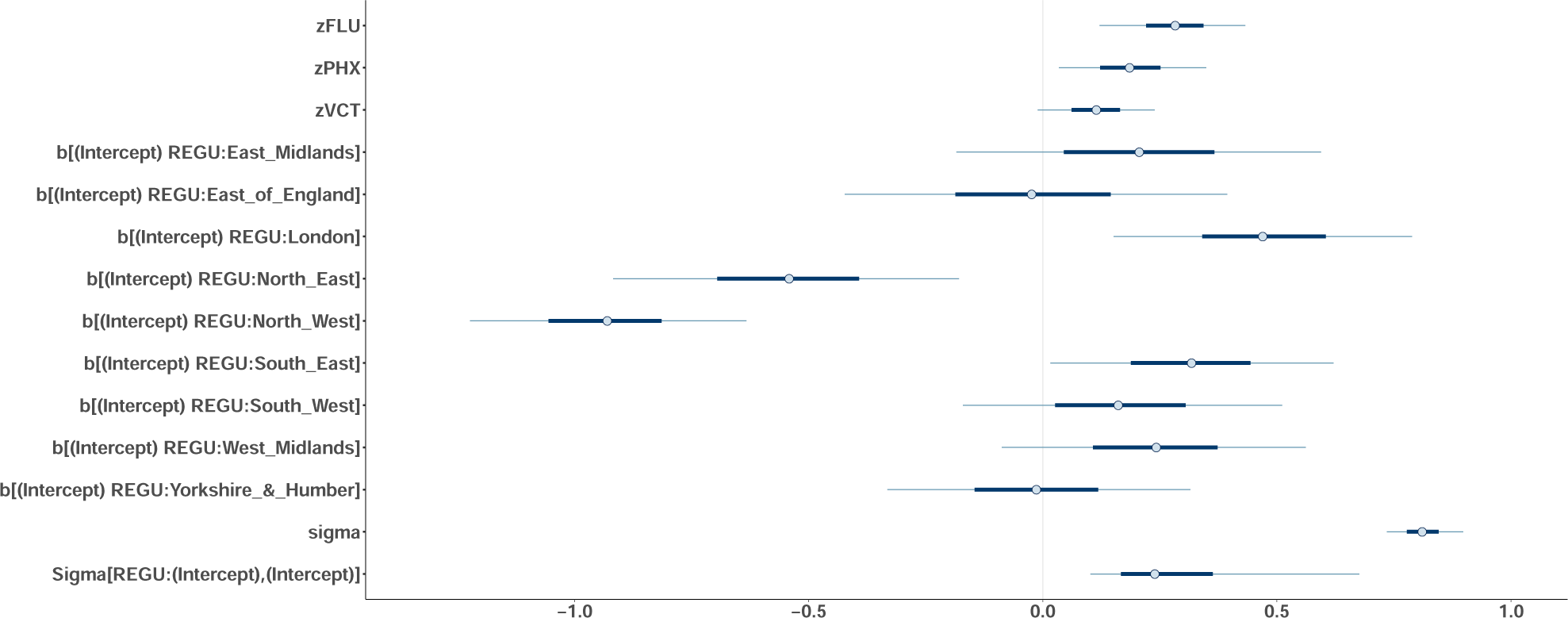
2nd stage model parameters and 90% credible intervals.

Table 2 shows the parameter estimates and 90% credible intervals after simulating (x100) the random effects from all four models (applied to the 3^rd^ Injection) before second stage modelling. The second stage model fits increasingly well as the first stage model improves.

**Table 2:** parameter estimates from 2^nd^ stage modelling of simulated random effects.

| | $\mathcal{A}$ | | $\mathcal{B}$ | | $\mathcal{C}$ | | $\mathcal{D}$ | |
| --- | --- | --- | --- | --- | --- | --- | --- | --- |
| Bayes $R^2$ | 0.174 | | 0.270 | | 0.336 | | 0.354 | |
|  | estimate | 90% credible | estimate | 90% credible | estimate | 90% credible | estimate | 90% credible |
| Flu vaccination | 0.217 | (0.046,0.386) | 0.219 | (0.049,0.390) | 0.302 | (0.142,0.461) | 0.280 | (0.119,0.442) |
| Public Health | 0.292 | (0.117,0.464) | 0.199 | (0.025,0.370) | 0.278 | (0.115,0.439) | 0.191 | (0.026,0.356) |
| Vaccination Centres | 0.123 | (-0.016,0.263) | 0.103 | (-0.033,0.238) | 0.149 | (0.020,0.278) | 0.109 | (-0.021,0.239) |
| East Midlands | 0.114 | (-0.202,0.486) | 0.217 | (-0.182,0.634) | 0.215 | (-0.168,0.611) | 0.203 | (-0.196,0.607) |
| East of England | -0.217 | (-0.653,0.113) | -0.110 | (-0.534,0.305) | -0.076 | (-0.483,0.324) | -0.007 | (-0.428,0.413) |
| London | 0.127 | (-0.142,0.442) | 0.397 | (0.052,0.754) | 0.288 | (-0.028,0.612) | 0.457 | (0.125,0.794) |
| North East | 0.039 | (-0.271,0.365) | -0.362 | (-0.765,0.020) | -0.324 | (-0.704,0.041) | -0.538 | (-0.937,-0.150) |
| North West | -0.356 | (-0.713,-0.012) | -0.742 | (-1.072,-0.407) | -0.790 | (-1.103,-0.471) | -0.905 | (-1.213,-0.593) |
| South East | -0.051 | (-0.338,0.218) | 0.161 | (-0.153,0.479) | 0.242 | (-0.058,0.547) | 0.307 | (0.000,0.617) |
| South West | 0.069 | (-0.222,0.388) | -0.002 | (-0.355,0.349) | 0.219 | (-0.116,0.564) | 0.160 | (-0.188,0.511) |
| West Midlands | 0.196 | (-0.090,0.551) | 0.279 | (-0.067,0.637) | 0.210 | (-0.119,0.545) | 0.233 | (-0.105,0.575) |
| Yorkshire & Humber | 0.122 | (-0.155,0.446) | 0.088 | (-0.250,0.427) | 0.062 | (-0.262,0.386) | -0.011 | (-0.345,0.321) |

Bayes R^2^ improves when second stage modelling is applied to the random effects obtained from better first stage models ***𝒟*** is preferred to other models by ML value and leads to higher Bayes R^2^). All four models lead to 2^nd^ stage models which show significant elevation (90% credible interval) for Flu vaccination and the rise in Public Health budgets as predictors of the simulated random effects, whilst the number of Vaccination Centres is only significant if the random effects are simulated from model ***𝒞***. North West Region is a significant negative predictor with all four models, and the North East is significant negative with model ***𝒟***. London is a significant positive predictor of random effects from models **ℬ** and ***𝒟***. The South East is borderline positive with model ***𝒟*** only.

A comparable table for the 1^st^ dose shows that Bayes R^2^ rises from 0.361 for modelling the estimated random effects from ***𝒜***, to 0.442 for ***ℬ*** 0.594 for ***𝒞***, and 0.593 for ***𝒟***. All four models lead to 2^nd^ stage models which show significant elevation (90% credible interval) for Flu vaccination and the rise in Public Health budgets as predictors of the simulated random effects, and likewise for London and South East regions, whilst North West is negative, and North East is negative for ***ℬ***, ***𝒞***, and ***𝒟***. South West is positive for ***𝒞*** and ***𝒟***.

For the 2^nd^ dose, Bayes R^2^ rises from 0.376 for modelling the estimated random effects from ***𝒜***, to 0.447 for B, 0.586 for ***𝒞***, and 0.587 for ***𝒟***. All four models lead to 2^nd^ stage models which show significant elevation (90% credible interval) for Flu vaccination and the rise in Public Health budgets as predictors of the simulated random effects, and likewise for London and South East regions, whilst North West is negative, and North East is negative for ***ℬ***, ***𝒞***, and ***𝒟***. South West is positive for ***𝒞***, and ***𝒟***, and borderline for ***𝒜***.

Chart 5 shows the scaled estimated random effects from model ***𝒟*** against the prediction from second stage modelling, for each dose.

**Chart 5:**
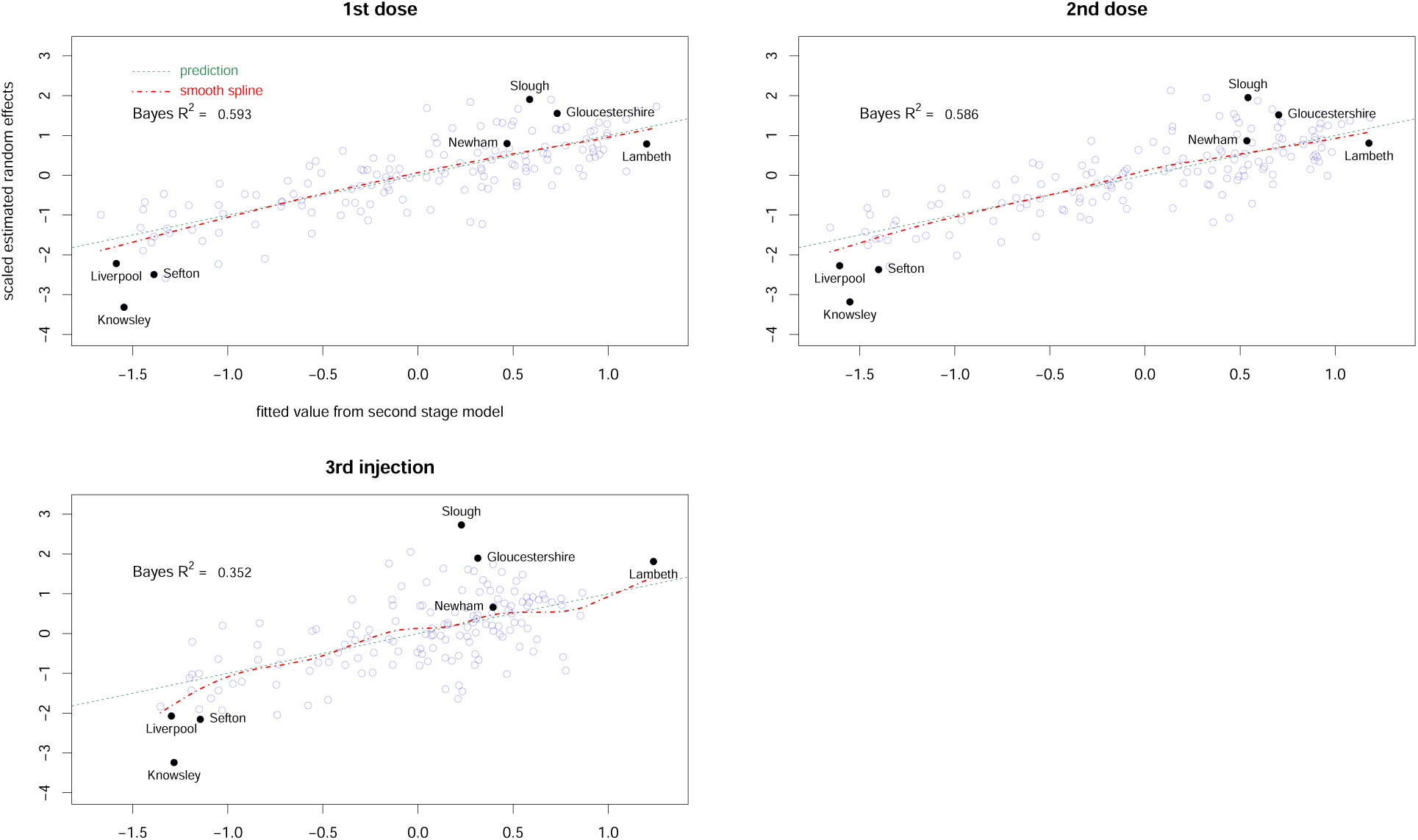
2^nd^ stage prediction of scaled estimated random effects from model *𝒟*.

## Discussion

There is extensive literature on Covid-19 vaccine uptake. Before the vaccines were deployed, a telephone and web survey of attitudes, considered by the government’s advisory group SAGE in December 2020 (see [4]) found “marked differences existed by ethnicity, with Black ethnic groups the most likely to be COVID-19 vaccine hesitant followed by the Pakistani/Bangladeshi group.

Other White ethnic groups (which includes Eastern European communities) also had higher levels of COVID-19 vaccine hesitancy than White UK/White Irish ethnicity”. SAGE cited previous studies to conclude “Barriers to vaccine uptake include perception of risk, low confidence in the vaccine, distrust, access barriers, inconvenience, socio-demographic context and lack of endorsement, lack of vaccine offer or lack of communication from trusted providers and community leaders.”

Later, a meta-analysis of international studies by Qiang Wang (see [3]) found that “Gender, educational level, influenza vaccination history, and trust in the government were strong predictors of COVID-19 vaccination willingness” and that healthworkers were less willing than the general public. A literature review of UK studies by Atiya Kamal (see [5]) concluded “Ethnic minority status was associated with higher vaccine hesitancy and lower vaccine uptake compared with White British groups. Barriers included pre-existing mistrust of formal services, lack of information about the vaccine’s safety, misinformation, inaccessible communications, and logistical issues”. A strategy to overcome vaccine hesitancy was advocated by Mohammed S Razai (see [8]), highlighting Confidence (importance, safety and efficacy of vaccines); Complacency (perception of low risk and low disease severity); Convenience (access issues dependent on the context, time and specific vaccine being offered); Communications (sources of information); and Context (sociodemographic characteristics). In an article entitled “What must be done to tackle vaccine hesitancy and barriers to COVID-19 vaccination in migrants?” (see [9]) Alison Crawshaw highlighted “mistrust of the state and health system, stemming from historical events, data sharing policies and dissatisfaction with the initial handling of the pandemic” and advocated “engaging with communities to understand their concerns or barriers to vaccination and working together to co-develop tailored approaches to encourage uptake and rebuild trust”.

A cohort analysis by Helen Curtis *et al*. (see [7]) of 57.9 million patients’ primary care records, concerns the period from 8 December 2020 to 17 March 2021. Of patients aged ≥80 years not in a care home (JCVI group 2) 94.7% received a vaccine, but with substantial variation by ethnicity (White 96.2%, Black 68.3%) and deprivation (least deprived 96.6%, most deprived 90.7%). Patients with pre-existing medical conditions were more likely to be vaccinated with two exceptions: severe mental illness (89.5%) and learning disability (91.4%).

The Office for National Statistics Coronavirus (COVID-19) latest insights: Vaccines, (as of 28 January 2022) ^32^ shows survey data on the proportion receiving 3 vaccinations by occupation groupings, ranging from 80.4% (Health) to 48.0% (Elementary trades and related occupations); and for more specific occupations; and by ethnicity, ranging from 68.4% (White British) to 33.9% (Black Caribbean).

An ONS technical article ^33^ explains the logistic regression models applied to vaccination status of individuals, controlling for sex, ethnicity, age, geographical region, urban or rural classification of their address, deprivation percentile, household size, whether the household was multigenerational. The model outputs ^34^ include regional variation, with negative impact of residence in the North West amongst persons aged 18 – 34.

An ONS study of ethnic contrasts in coronavirus death rates published on 26 January 2022 ^35^ found that “Location, measures of disadvantage, occupation, living arrangements, pre-existing health conditions and vaccination status accounted for a large proportion of the excess rate of death involving COVID-19 in most ethnic minority groups; however, the Bangladeshi ethnic group and men from the Pakistani ethnic group remained at higher risk than White British people in the third wave, even after adjusting for vaccination status”.

A 2018 study of childhood immunisation rates in Italy by Veronica Toffolutti ^36^, found that reductions in public health budgets were a significant predictor of falling immunisation rates.

In the geographical analysis reported here, I initially sought to learn whether the current very low booster uptake in particular small areas was unexpected, or could be predicted from the national data on the UK Coronavirus Dashboard, at Middle Super Output level.

In each MSOA, the data shows the number of people eligible for the vaccine, and the cumulative number actually vaccinated with the first and second doses, and the 3^rd^ Injection. The latter covers both people receiving the booster, and those with compromised immunity who were given a 3^rd^ primary dose. The ratio of those vaccinated to those eligible, is the uptake. But the actual numbers vaccinated and eligible give more information, and can be modelled as a binomial variable.

The Generalized Additive Model, implemented in R with the mgcv package, is an established technique for non-linear modelling. The variance in this data exceeds what would be expected for a binomial variable, but can be estimated by relaxing the “binomial” assumption to “quasibinomial”. Using demographic, employment and local health data as predictors, such models fit this data well.

The models are improved by including a random effect assigned to the Upper Tier Local Authority within which the MSOA is found, allowing other aspects of the UTLA to influence the prediction.

Four different models for each of the three vaccinations, all gave a good fit to the data, with the proportion of deviance explained ranging from 95.4% to 97.2%. Chart 1 shows how well the preferred model **𝒟** performs for the 3^rd^ injection. This close fit means that the observed values in a particular MSOA are almost always near the prediction from the national data.

The covariate structure was simplified to enable models to be computable on a PC. For example, whilst population data is available for each year of age, the models use broader categories such as aged 15-24 or 25-44. Even so, **𝒟** has 33 smoothers, each with a smoothing parameter which must be estimated during fitting, along with 341 coefficients.

The smoothers for **𝒟**, some of which are shown in Chart 2, show the effect of individual covariates in the context of all others in the model. The most powerful predictors are the Index of Multiple Deprivation, the population proportions aged 15-24, aged 25-44, of Other White ethnicity, South Asian (Indian, Pakistani and Bangladeshi combined), and the combined group of African, Caribbean, Other Black, and Other ethnicity.

The negative slope of the first smoother means that a higher IMD leads to a lower predicted uptake if all other variables are unchanged. Education Deprivation (a specific domain within the overall index IMD) also has a negative slope, although its effect is smaller (shown on Chart 2 with “narrow scale”). All six most powerful predictors have smoothers with negative slope. The dependence on age is to be expected, as the vaccine rollout targeted different age groups at different times.

Ethnicity has significant impact on uptake, as many other studies have found. The smoother for the “Other White” group has the widest range, comparable to that for IMD, showing that the “Other White” proportion of the population has a strong impact on predicted uptake.

Several other smoothers have small positive slope, such as those for employment in Public Adminstration, or Health and Social Care. The latter indicates that despite the well publicised vaccine hesitancy amongst a minority of NHS and care staff, the overall impact of employment in these sectors is to increase the uptake of the 3^rd^ Injection. This is consistent with the ONS data (see [32]). For the 1^st^ and 2^nd^ doses, the increase is clear only when the proportion of Health and Care staff exceeds the 20^th^ percentile of this covariate.

The smoother for average distance to A&E also has a positive slope, suggesting that people living further from an emergency department may be more concerned to take the vaccination. Likewise, increasing Health Deprivation is associated with increasing uptake. Conversely, increased GP registration is associated with decreasing uptake. However, there is some uncertainty in the GP registration data, and NHS Digital have commented on the excess of GP registrations over ONS population estimates.^37^

Fitting any of these models gives estimates for the effect of each of the demographic, employment, and health variables included as predictors, and the estimated “random effect” of each of the 149 UTLA. For example “Knowsley” has a strong negative impact, and “Slough” has a strong positive impact. These effects are not to be confused with the actual uptake of vaccination in the local authority, which may be low or high due to deprivation or other fixed effects, and which may vary widely amongst MSOAs within the local authority. Uptake of the 3^rd^ Injection varied from 32.2% to 57.6% within Knowsley, from 17.3% to 65.5% in Liverpool, and from 36.9% to 75.8% in Sefton.

The random effect alters the prediction from the fixed effects alone. If Beckton Park (in Newham) had been located in Slough with identical demography, employment, and health indicators, the predicted uptake of the 3^rd^ Injection would rise by 18%, whilst if it were in Knowsley the prediction would fall by 29%.

The range in magnitude of the random effects is comparable to that of ethnicity groups. On the scale of the linear predictor, the smoothers for Other White, South Asian, and Afro-Caribbean groups range from 0.09 to -0.70, 0.05 to -0.42, and 0.07 to -0.49 respectively, each smoother descending as the ethnicity proportion rises. The random effects range from -0.15 (Knowsley) to 0.50 (Slough).

The difference between the random effects for Knowsley and Slough is greater than the maximal difference between any two MSOA due to the population proportion of South Asian or Afro-Caribbean ethnicity, and only slightly less than the maximal difference due to the proportion of Other White.

The second stage of modelling focused on the 149 random effects, seeking to explain their estimated values in terms of other information at UTLA level. This stage tested the impact of flu vaccination rates, public health budgets, vaccination centres, and Region, treated as a random factor. Ideally all of the UTLA and MSOA level variables would be incorporated in a single model. But that appeared prohibitively slow to compute, so the random effects estimates from the first stage mgcv modelling were considered as observations, to be modelled in their own right. Simulated random effects generated from the first stage model were used to refine the parameter estimates and credible intervals from the second stage, but this made little difference. For example the parameters and credible intervals for model **𝒟** in Chart 4 are similar to those found for **𝒟** after simulation in Table 2.

The second stage model passes Bayesian checks and shows clear effects of Region, flu vaccination, and public health budget increase. Chart 5 shows the extent to which the chosen predictors actually explain the estimated random effects. None of the points are outliers (by pareto_k) and the model fits well for the 1^st^ and 2^nd^ doses with Bayes R^2^ ∼ 0.6. The lower value for the 3^rd^ injection, ∼ 0.35, suggests that there may be other relevant covariates at UTLA level.

It is striking that Knowsley, Liverpool and Sefton all appear at the lower left of Chart 5 and Knowsley is conspicuously low for all three vaccinations. The Merseyside local authorities are amongst the most deprived in England, but IMD already appears in the first stage model **𝒟** as a fixed effect so was not expected to have any impact on the second stage model. Indeed, if IMD is averaged over the MSOA within each UTLA and then used as a predictor in second stage modelling, it has no significant effect.

Simply being in the North West is the most powerful predictor of low uptake in this model, and in addition Knowsley had the 13^th^ lowest increase in public health funding (and the 3^rd^ lowest within the North West), rising by only 0.88%. Newham benefits from being in London and from the larger increase in its public health budget (rising by 2.09%), and possibly from the vaccination centre located in Olympic Park, an outlier with positive residual (see Chart 1). Flu vaccination rates are higher in Knowsley (49.2) than in Newham (45.5), so cannot explain the disparity in random effects.

Slough benefits from being in the South East and possibly from a vaccination centre, whilst its flu vaccination rate and rise in public health budget are both close to their respective mean values.

However the random effects for Slough also exceed prediction. The highest predicted value is in Lambeth, where the flu vaccination rate is below average, but which benefits from being in London with a 4.88% rise in public health budget and 3 vaccination centres. Its estimated random effect is close to prediction for all 3 doses.

All such conclusions depend on the validity of the models. The first stage MSOA level modelling fits very well, and the resulting estimated random effects from all four models are highly correlated. That is, the random effects are not simply artefacts of the model, once ethnicity is included. The estimated random effects change dramatically if ethnicity is omitted from the model.

The UTLA level effects could be described as a postcode lottery, as they are not explained by the population characteristics controlled for in the fixed effects, but are associated with other geographical factors. However, a “lottery” suggests pure chance whereas economic policy decisions affect Regional disparity and public health budgets, which in turn affect Covid vaccination rates.

Much of the literature has focused on “vaccine hesitancy” of specific population subgroups. The modelling here confirms the impact of ethnicity along with deprivation and age, but also identifies additional factors, characteristic of the locality rather than the population living in it, consistent with evidence on childhood immunisations in Italy.

These models indicate that whatever other barriers exist due to deprivation and within particular ethnicities, the annual change in local authority public health budgets is also a significant factor. Therefore, increasing local public health allocations would be one simple way to improve Covid vaccine uptake.

## Data Availability

All data produced in the present study are available upon reasonable request to the authors

## Acknowledgements

Thanks to David Taylor Robinson for helpful comments including the AHAH dataset and the suggestion that budgets could be a predictor, and to Isabelle Whelan for an unpublished essay “Austerity, NHS reform and vaccine uptake”, written before the pandemic.

